# Splitting ratio in sevoflurane vapouriser-a relook on calculations

**DOI:** 10.1101/2024.04.11.24305499

**Authors:** D Girijanandan Menon, Manjit George, Salih V Salim, R. Sreekumar, Sam Philip, Roshin K Mathew, Princymol Bju, Merin Helen Mathai

**Author notes:** <u>Contribution by authors</u> (author name with abbreviation of name is given in brackets) Manjit George (MG) Salih V Salim (SS) D Girijanandan Menon (DG) R.Sreekumar (RS) Sam Philip (SP) Roshin K Mathew (RM) Princymol Bju (PB) Merin Helen Mathai (MM) DG is involved in concept development, derivation of formula, writing up the manuscript, editing, concept validation and reliability of the concept as teaching aid in the department. MG Mainlead in concept validation, manuscript editing, verified thereliability of the concept as teaching aid in his instituion SS concept development, concept proof verification, manuscript editing, verified the reliability of the concept as teaching aid in his instituion RS did the derivation styling, illustrations and supplementary file editing SP involved in concept validation, and verification by department faculty, granted permission for including the explanation in discussions on vapouriser, ultiisation of concept for post graduate teaching in the Department of Anaesthesiology RM,PB&MM are non author contributors for implementation of the concept for postgraduate teaching and for analyzing the reliability of the concept as a better teaching aid in the Department of Anaesthesiology.

## Abstract

Textbooks of anaesthesia describe in detail the calculations associated with splitting ratios created in *variable bypass vapourisers*. These calculations are based on assumptions of carrier gas flow (eg:assume 150 ml) as practised with *measured flow vaporizers*[1,2]. Carrier gas flow in a variable bypass vaporizer is agent specific,dynamic,unknown and operator independent.Therefore assuming carrier gas flow in calculations related to variable bypass vaporisers is without rationale[3]. In spite of technological advance,the amount of vapour produced is calculated based on clinically impractical assumptions. A simple method and formula is suggested for the quick estimate of vapour output, carrier gas flow and determine the splitting ratio while operating a sevoflurane variable bypass vaporizer.

## Introduction

### What is the current method to estimate the amount of vapour produced in vapourising chamber?

Carrier gas flow is assumed to estimate the amount of vapour produced in vapourising chamber.

### What is the new method suggested to calculate the amount of vapour produced in vapourisng chamber?

Vapour produced in vaporising chamber is simply the product of the percentage of the concentration control dial setting and the fresh gas flow in ml per minute. Instead of assuming the carrier gas flow, it is calculated real time from the settings. An attempt is made to simplify the calculations by logically relating the assumptions to the operator’s act of turning the vaporizer dial to achieve the required output concentration in a variable bypass vapouriser. This will enable judicious control of the concentration of the volatile requirements like achieving the ED_95_,uptake by circulation,tissues and the priming dose of vapour needed for ventilation etc in low flow anaesthesia[4]. Therefore the volume of vapour produced in the vaporising chamber of a modern variable bypass vaporiser can be estimated instantaneously and achieve the targeted amount of vapour proportional to the fresh gas flow and dial setting.

#### Proposed explanation

The dial settings of the variable bypass vaporizers currently used are off, zero and the markings in percentage. When the vapouriser dial is in the off position fresh gas flow does not enter the vapouriser.It exits via the back bar of the anaesthesia machine. In the zero position all the fresh gas flow pass through the bypass chamber of the vapouriser manifold. When the dial is set to c%, the fresh gas flow (f) splits into two streams namely, the vapourising chamber(carrier gas) flow(x) and bypass flow(f-x)as explained in the Figure1.

**Figure 1.**
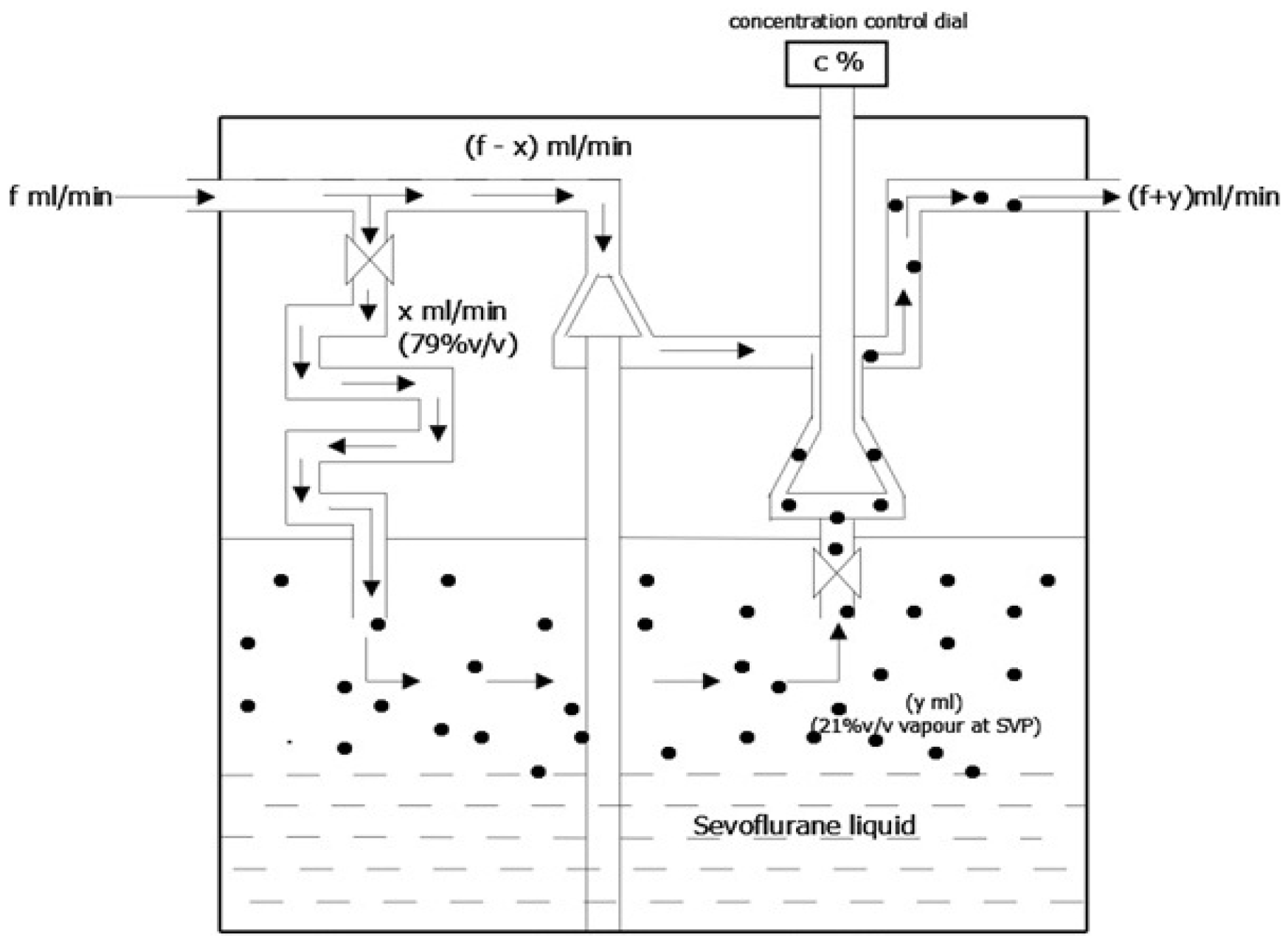
showing line diagram of cross section of temperature compensated variable bypass vapouriser with baffles.Valves are denoted by the X shape. Cones represent the concentration control and the thermostat,c -dialled concentration of the volatile agent, f-fresh gas flow entering vapouriser inlet, x-the carrier gas flow and y-the volume of vapour produced inside the vapourising chamber, v-the volume & SVP-the saturated vapour pressure.Black dots represent the vapour molecules and arrows represent the dirction of flow of fresh gas.

The carrier gas flowing above the volatile liquid in the vapourising chamber is enriched by the saturated vapour volume(y ml). This is equal to the dialled in percentage concentration of the vaporizer multiplied by the fresh gas flow(f ml/min)entering the vaporizer inlet. The vapour so produced has a saturated vapour pressure of 160 mm of Hg at 20°C at atmospheric pressure. Therefore as per Dalton’s law of partial pressure,concentration of saturated vapour produced is 160 mmHg/760mmHg=0.21(21% volume) which is lethal for humans. Carrier gas forms the remaining 79%(600/760) volume(x ml) of the mandatory atmosphere above the volatile liquid.When the fresh gas flow(f) is 1000 ml per minute with a vaporizer dial setting(c) of one percent, the volume of vapour added to the carrier gas flow is simply 10 ml(y ml). The *vapourising chamber outpu*t concentration of 21% sevoflurane vapour (in a volume of ten ml) in the carrier gas flow of unknown volume (*x* ml) is reduced to the clinically safe concentration of one percent vapour at the *vapouriser output*. This occurs on dilution with bypass gas flow(f-x) at the vaporizer outlet. The carrier gas flow (x ml/min) may be calculated from the equation x/y=(600/760)/(160/760)=x(ml/min)/ 10 (ml/min) ie x=3.75*10=38 ml. Therefore the bypass flow is 1000-38=962 ml and the splitting ratio is 962/38=25, ie 25:1. The carrier gas flows for a set of dial settings and fresh gas flows are given in the Table 1 below.

**Table 1.**
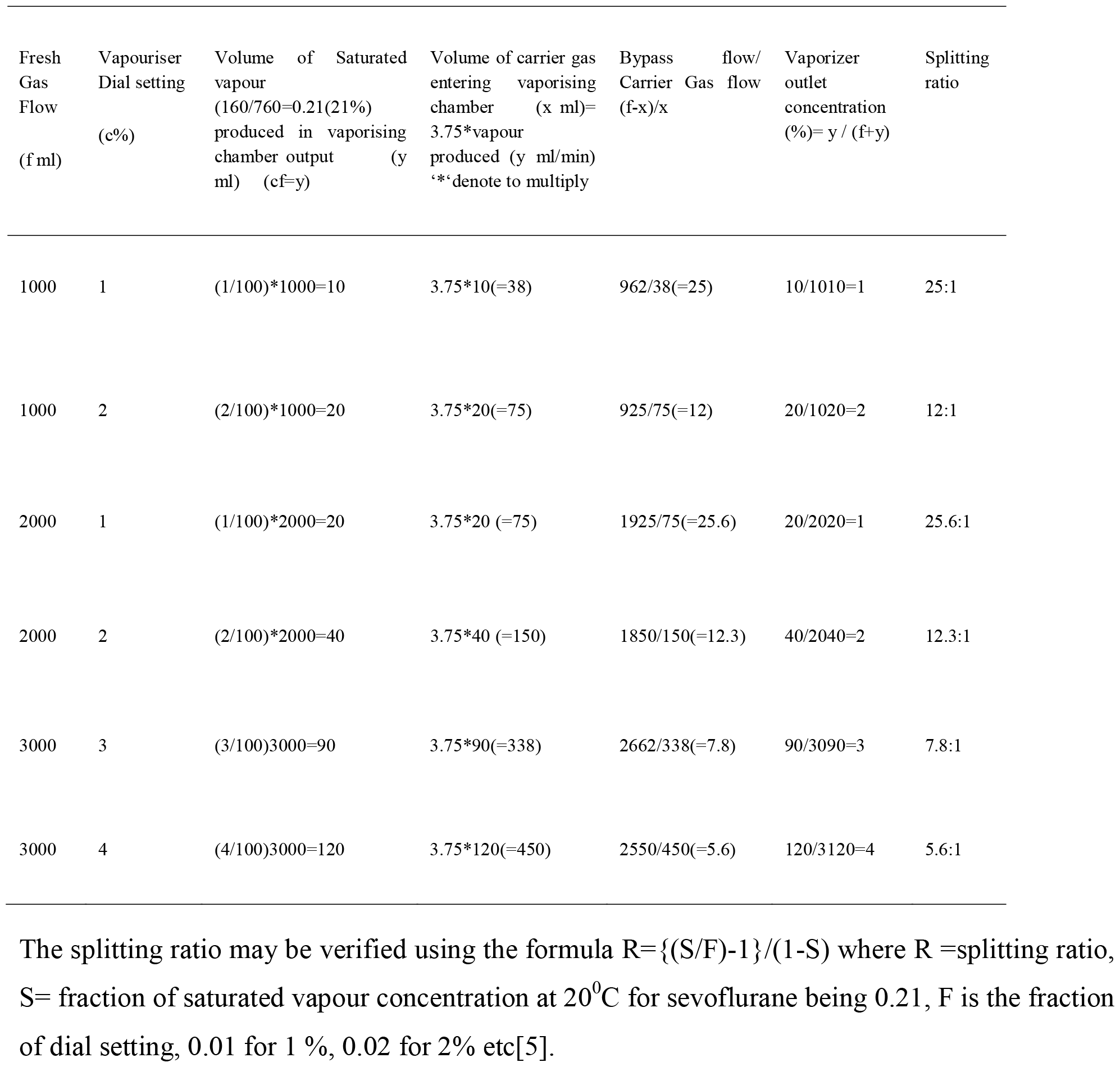
Calculation of vapour output, bypass & carrier gas flow,vaporiser outlet concentration and splitting ratio of a variable bypass sevoflurane vapouriser.3.75 is the ratio of partial pressure of carrier gas(600mm Hg)at atmospheric pressure(760mm Hg) to the saturated vapour pressure of sevoflurane(160mmHg)at 20°C produced in the vapourising chamber.

Simplified formula for calculating the splitting ratio in a sevoflurane variable bypass vaporizer is given below. Please see the appendix for derivation of the formula. S=(1/3.75c)-1, where S is splitting ratio,c is fraction of the dialled concentration of sevoflurane (c = 0.01 for dial setting of 1 %). Constant 3.75 is the ratio of partial pressure of carrier gas at atmospheric pressure to the saturated vapour pressure of sevoflurane vapor produced in the vapourisng chamber at 20 ° C.

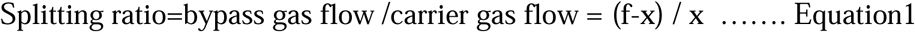

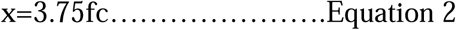

On applying Equation 2, Equation 1 becomes

Splitting ratio=(f-3.75fc) / 3.75f=f(1-3.75c) / 3.75fc =(1-3.75c) / 3.75c

Splitting ratio= (1/3.75c) -1

When dialled concentration is 1%, c=0.01,splitting ratio=(1/3.75*0.01)-1=26.66 -1=25.66 Therefore splitting ratio is approximately 25:1

## Conclusion

Reliable estimation of the amount of anaesthetic vapour produced in the vapourising chamber andthat delivered at the the vaporizer outlet is clinically more important for the operator while providing inhalational general anaesthesia. Calculating the same based on assumptions of carrier gas flow is not logical. The above explanation helps in the safer use of the volatile agent during low flow anaesthesia. Vapour out put is the product of fresh gas flow and set dial concentration. Carrier gas flow for sevoflurane vaporizer is 3.75 times the vapour output. Splitting ratio derived above is independent of fresh gas flow but dependent on the dialled concentration of the agent used.

## Data Availability

All data produced in the present work are contained in the manuscript

## Appendix

### Derivation of splitting ratio of a modern sevoflurane vapouriser

Saturated vapour pressure (SVP) of sevoflurane at 20°C = 160 mm Hg Atmospheric Pressure= 760mm Hg

Partial pressure of carrier gas in vapourising chamber =760-160 =600mm Hg

Concentration of sevoflurane vapour in vapourising chamber= 160760=0.21(21%)

Concentration of carrier gas(C_cg_)in vapourising chamber =600760=0.79(79%)

f = fresh gas flow entering the vapouriser inlet (ml/min)

c = concentration in percentage or fraction dialed on the concentration control dial (1%=0.01)

x = carrier gas volume entering the vapouring chamber (ml/min)

y = vapour produced in the vapourising chamber (ml/min)

f-x = bypass gas flow (ml/min)

x + y = sum of the volumes of carrier gas and vapour (ml/min)

At 20°C, y is 21 % of vapourising chamber atmosphere and x is 79% in a modern sevoflurane vapouriser. Therefore the ratio between carrier gas and vapour produced is given by

x / y=(79%) / (21%)= (600/760) / (160/760)=600 / 160=3.75

So x=3.75 *y and y=f*c

Therefore x=3.75*f*c

When the flow split in the variable bypass vapouriser occurs at the vapouriser in let and outlet the splitting ratio of the fresh gas flow is given by S=(f-x) / x & S= (f-x) / (x+y) respectively. By applying the values of x & y as given above the splitting ratio formulae may be derived as given below. The splitting ratio(s) for effective gas split at vapouriser inlet is given by s = (1/3.75*c)-1 and the exit flow split occuring at the vapourising chamber outlet is s=(1/4.75*c) – Ccg, where c is the concentration control dial setting and Ccg_g_ is the carrier gas concentration in percentage. Thus it is proof that splitting ratio is independent of fresh gas flow. It depends on the dialled concentration and the constant specific for the volatile anaesthetic (sevoflurane). This constant is the ratio of the partial pressure of carrier gas at atmospheric pressure to the saturated vapour pressure of the vapour produced in the vapourising chamber generally at 20°C. This constant depends on the concentration of the vapour produced inside the vapourising chamber. For halothane this constant is (760-243) / 243=2.1.

## Reference

1. Miller RD, Michael P. Bokoch, Stephen D. Weston. MIller’s Anaesthesia. 9th ed. 2020.

2. Jan Ehrenwerth, Ehrenwerth JB, Berry JM. Anesthesia Vaporizers, Chapter3, page 68-70. In: Anaesthesia Equipment, Principles & Practice. 2nd ed. Elsevier; p. 68–70.

3. Dhulkhed V, Shetti A, Naik S, Dhulkhed P. Vapourisers: Physical Principles and Classification. Indian J Anaesth [Internet]. 2013 [cited 2024 Apr 5];57(5):455–63. Available from: https://www.ncbi.nlm.nih.gov/pmc/articles/PMC3821262/

4. Jacobs D. Closed circle anesthesia [Internet]. NYSORA. 2023 [cited 2024 Mar 6]. Available from: https://www.nysora.com/closed-circle-anesthesia/

5. Leigh JM. Variations on a theme: splitting ratio. Anaesthesia. 1985 Jan;40(1):70–2.

